# Genetic immune response and antibody repertoire of heterologous ChAdOx1-BNT162b2 vaccination in a Korean cohort

**DOI:** 10.1101/2022.02.07.22270617

**Authors:** Hye Kyung Lee, Jinyoung Go, Heungsup Sung, Seong Who Kim, Mary Walter, Ludwig Knabl, Priscilla Furth, Lothar Hennighausen, Jin Won Huh

**Affiliations:** National Institute of Diabetes, Digestive and Kidney Diseases, National Institutes of Health, Bethesda, MD 20892, USA; Department of Biochemistry and Molecular Biology, Asan Medical Center, University of Ulsan College of Medicine, Seoul, Korea; Department of Laboratory Medicine, Asan Medical Center, University of Ulsan College of Medicine, Seoul, Korea; Clinical Core, National Institute of Diabetes, Digestive and Kidney Diseases, Bethesda, MD 20892, USA; TyrolPath, Zams, Austria; Departments of Oncology & Medicine, Georgetown University, Washington, DC, USA; Department of Pulmonary and Critical Care Medicine, Asan Medical Center, University of Ulsan College of Medicine, Seoul, Korea

## Abstract

Heterologous ChAdOx1-BNT162b2 vaccination induces a stronger immune response than two doses of BNT162b2 or ChAdOx1. Yet, the molecular transcriptome, the germline allelic variants of immunoglobulin loci and anti-Omicron antibody levels induced by the heterologous vaccination have not been formally investigated. Moreover, there is a paucity of COVID vaccine studies including diverse genetic populations. Here, we show a robust molecular immune transcriptome and antibody repertoire in 51 office workers from the Republic of Korea after a heterologous ChAdOx1-BNT162b2 vaccination or a homologous ChAdOx1-ChAdOx1 vaccination. Anti-spike-specific IgG antibody levels in the heterologous group increased from 14,000 U/ml to 142,000 AU/ml within eight days after the BNT162b2 vaccination. In contrast, antibody levels in the homologous group increased two-fold after the second ChAdOx1 dose. Antibody titers against the Omicron spike protein as compared to the ancestral strain were reduced to a lesser extent in the heterologous group. RNA-seq conducted on immune cells demonstrated a stronger activation of interferon-induced genetic programs in the heterologous cohort. An increase of specific IGHV clonal transcripts encoding neutralizing antibodies was preferentially detected in the heterologous cohort. Enrichment of B cell and CD4+ T cell responses were observed following both heterologous and homologous vaccination using scRNA-seq, but clonally expanded memory B cells were relatively stronger in the ChAdOx1-BNT162b2 cohort. In summary, a heterologous vaccination with ChAdOx1 followed by BNT162b2 provides an innate and adaptive immune response exceeding that seen in homologous ChAdOx1 vaccinations but equivalent to that seen in homologous BNT162b2 vaccination.

## Introduction

The ChAdOx1 nCoV-19 vector (AZD1222) and BNT162b2 mRNA vaccines (hereafter referred to as ChAd and BNT, respectively) have been widely used and shown to induce robust immune responses against the spike protein of SARS-CoV-2. Both vaccines have shown remarkable efficacy in preventing COVID-19 disease. Based on the approval by government agencies and available supply, ChAd was the main vaccine used in the Republic of Korea (South Korea) in the spring of 2021. The manifestation of rare events, including thrombosis and thrombocytopenia syndrome, associated with adenovirus-based vaccines prompted pausing of the distribution of the ChAd vaccine in many countries, including South Korea^1^. This promoted partially vaccinated people to complete their vaccination with an mRNA vaccine, Pfizer/BioNTech (BNT) or Moderna. The ChAd– BNT heterologous vaccination strategy resulted in significantly greater immunoglobulin G (IgG) immune responses aimed against the SARS-CoV-2 spike protein compared to the ChAd-ChAd strategy^2-7^. While the immune response to different vaccine regimen has been investigated, there is limited knowledge about the molecular transcriptome responses, elicited by heterologous vaccinations. Here we hypothesized that the heterologous vaccination elicits unique molecular responses in immune cells by activating transcription of specific germline allelic variants. To test this hypothesis, we investigated immunogenicity and reactogenicity of two vaccine regimens, the heterologous ChAd-BNT and the homologous ChAd-ChAd.

While many vaccine studies are based on individuals with European and African ancestry^8,9^, an inclusion of samples from other genetic backgrounds would enable the equitable advancement of genomic vaccinology. In particular, there is a dearth of genomic vaccinology studies on Asian populations^10^. Addressing the distinct usage of germline allelic variants of immunoglobulin loci upon COVID-19 vaccination of different population might enhance our understanding of antibody production. Although there have been COVID-19 vaccine studies of Asian/Korean populations^11-14^, none of them had investigated the molecular immune response.

Here, we use serology and transcriptome analyses to investigate the immune response of office and laboratory workers from a major health care center in the Republic of Korea. Study participants received one ChAd dose followed either by a second ChAd dose or the BNT vaccine. The sequencing depth in this study, and our previous study on individuals receiving two doses of BNT facilitated the identification of an expanded immune response in individuals receiving the heterologous vaccination.

## Results

### Study design

A current discussion centers around the immune response elicited by heterologous ChAdOx1-BNT162b2 (referred to ChAd and BNT throughout the manuscript) vaccinations as compared to homologous two ChAd or two BNT doses. Here we compared the molecular immune response elicited by a heterologous vaccine regimen, ChAd followed by BNT, and a homologous two-dose ChAd regimen. Employees (n=51 (44 Females, 7 Males), median 38 years old, median BMI 22.4) from the Asan Medical Center, Seoul, Republic of Korea received the ChAd vaccine in June 2021 (Figure 1A-C; Supplementary Table 1). The study had been originally designed to investigate the immune response after two doses of ChAd and all 51 participants received the first ChAd dose in June 2021. Due to a revised vaccination policy 47 participants received the BNT vaccine as the second dose in August 2021. Four individuals still received the second ChAd dose. Four participants had underlying health conditions and two reported mild post-vaccine symptoms after the initial ChAdOx1 primary dose (Supplementary Table 1). We measured anti-spike IgG antibody levels, circulating cytokines levels and immune transcriptomes (RNA-seq) on peripheral immune cells prior to the vaccination and on days 2-4 (mean 2.8, median 3.0 days) and 7-10 (mean 7.9, median 7.0 days) post vaccination (referred to day 3 and day 8 throughout the manuscript). For selected samples we also conducted scRNA-seq.

**Figure 1.**
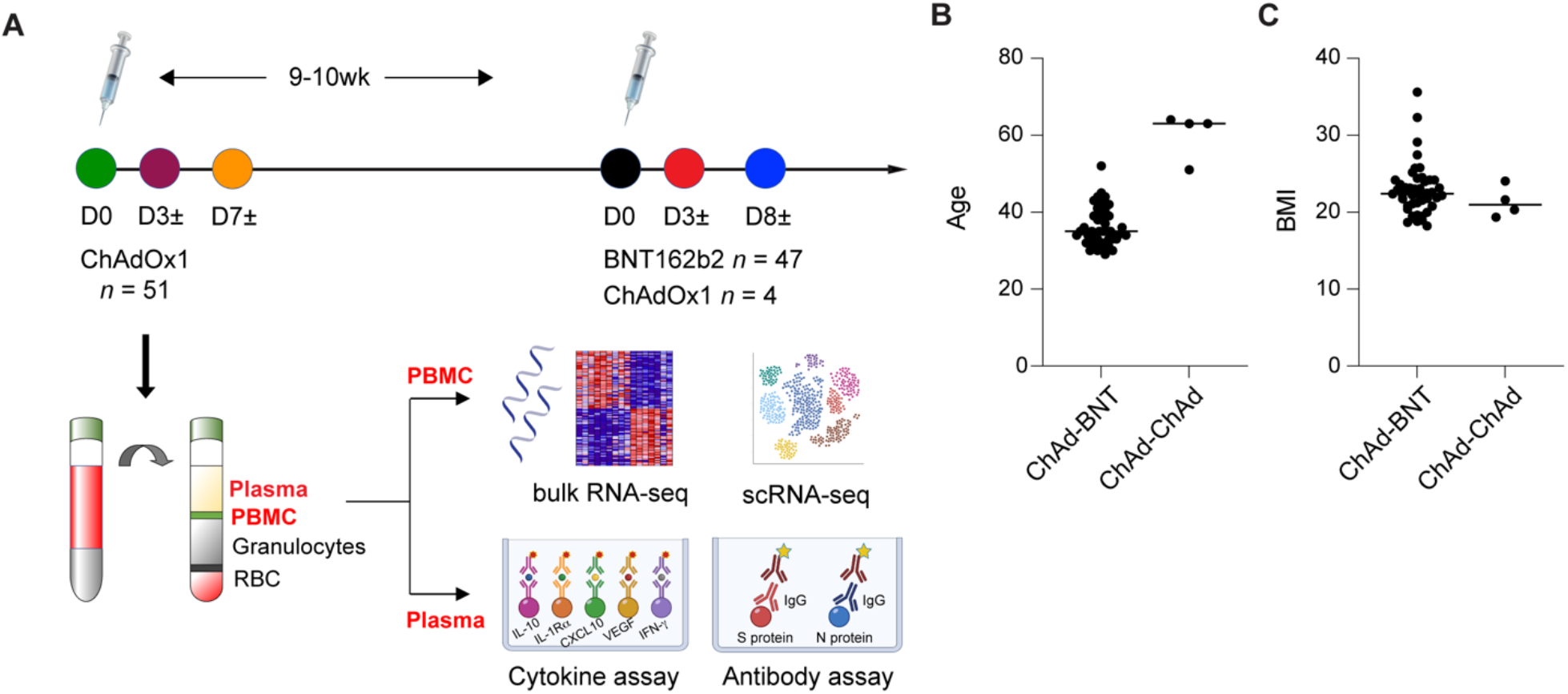
Experimental design. **A** Schematic presentation of the experimental workflow. All 51 study subjects received ChAd as a first vaccine dose, 47 received BNT as a second dose and 4 received ChAd as the second dose. Blood was collected on the day of vaccination (D0) and at day 3 (D3±) and day 8 (D8±) post vaccination as indicated by the colored circles. **B-C** Age and BMI distribution of participants in each study group. Further information is listed in Supplementary Table 1. Line at median.

### Antibody response after the first and second vaccination

First, we measured circulating antibody responses in plasma samples from 45 individuals receiving the heterologous ChAd-BNT vaccine regimen using enzyme-linked immunosorbent assay (ELISA) (Figure 2). On average, anti-spike (WH04) IgG levels were approximately 70 AU/ml within three days following the initial ChAd dose and 14,000 AU/ml within three days following the BNT vaccine dose (Figure 2A; Supplementary Table 2). Another 10-fold increase was observed at day 8 following the BNT vaccination. In contrast, no further increase in anti-spike IgG levels was observed in the homologous group between day 3 and 8 following the second ChAd dose (Figure 2B; Supplementary Table 2). Similar patterns were observed for the spike proteins from the Alpha, Beta, Gamma, Iota, Kappa, Zeta and Delta variants. Antibody concentrations against Omicron were reduced by approximately 70% in the ChAd-BNT group and 75% in the ChAd-ChAd group compared to the level from the ancestral strain (Figure 2A-C; Supplementary Figure 1). Previously we investigated the molecular immune response in a cohort receiving a two-dose homologous BNT-BNT vaccine regimen^15^ and the anti-spike IgG induction was in general higher in the heterologous group after the BNT dose as compared to the BNT-BNT homologous group (Supplementary Figure 1). Notably, the antibody titer against the Omicron variant was significantly higher in the heterologous group as compared to the homologous group (Supplementary Figure 1B).

**Figure 2.**
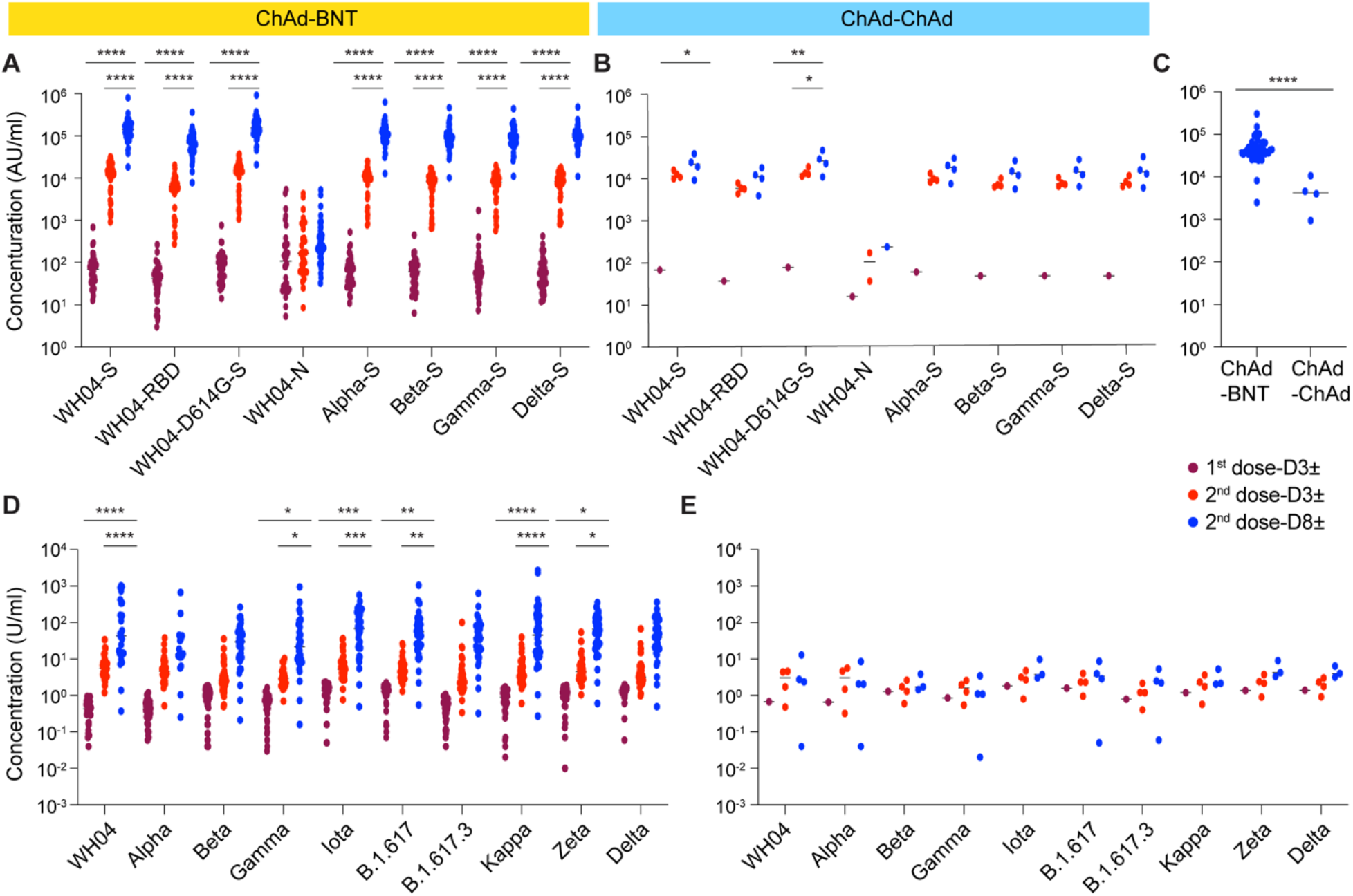
Antibody. **A-B** Plasma IgG antibody binding the SARS-CoV-2 RBD (spike) from different strains and the SARS-CoV-2 N protein in the ChAd-BNT and ChAd-ChAd vaccination groups. *p*-value are from 2-way ANOVA with Tukey’s multiple comparisons test. **C** Plasma IgG antibody binding the Omicron spike protein in the ChAd-BNT and ChAd-ChAd vaccination groups. **D-E** Neutralizing antibody response to virus spike protein of SARS-CoV-2 original and variants. *p*-value are from 2-way ANOVA with Tukey’s multiple comparisons test. \**p* < 0.05, \*\**p* < 0.01, \*\*\**p* < 0.001, \*\*\*\**p* < 0.0001. The individual’s data are listed in Supplementary Table 2. Line at median.

At this point in the pandemic, a critical question is whether antibodies induced by vaccination can neutralize current variants. Here, we measured the *in vitro* neutralization activity against different variants, including Delta (B.1.617.2) and used the degree of serum antibodies blocking the binding of ACE2 to SARS-CoV-2 variants’ spike as a proxy. While neutralizing activity for the ancestral strain and other variants increased between days 3 and 8 after the BNT162b vaccination in the heterologous cohort, no further increase was detected in the homologous cohort after the second ChAd dose (Figure 2D-E, Supplementary Table 2). We also found increased neutralizing activity against Delta variant in the BNT-BNT cohort^15^.

### Immediate immune response

Early responses to vaccination materialize in elevated levels of interferons and other cytokines. To gauge the early vaccine response, we measured serum levels of a panel of cytokines in all individuals prior to and after the initial ChAd vaccination and the second dose, either BNT or ChAd (Figure 3; Supplementary Table 3). Out of the 23 cytokines measured, a transient increase of circulating IL-10 and CXCL10 was observed within three days after the first vaccination and returned to baseline by day 7 (Figure 3A-B). Upon delivery of the second vaccine dose, CXCL10, IL-10 and IL-1Rα levels were statistically elevated by BNT but not by ChAd (Figure 3C-F). Only IL-1Rα was induced in both groups (Figure 3E-F). CXCL10 (IP-10) expression is rapidly activated following vaccination and viral infections and it has been described as a biomarker associated with COVID-19 severity^16-18^. Its regulation by IFN-*γ* is mediated by the JAK-STAT signaling pathway^19^.

**Figure 3.**
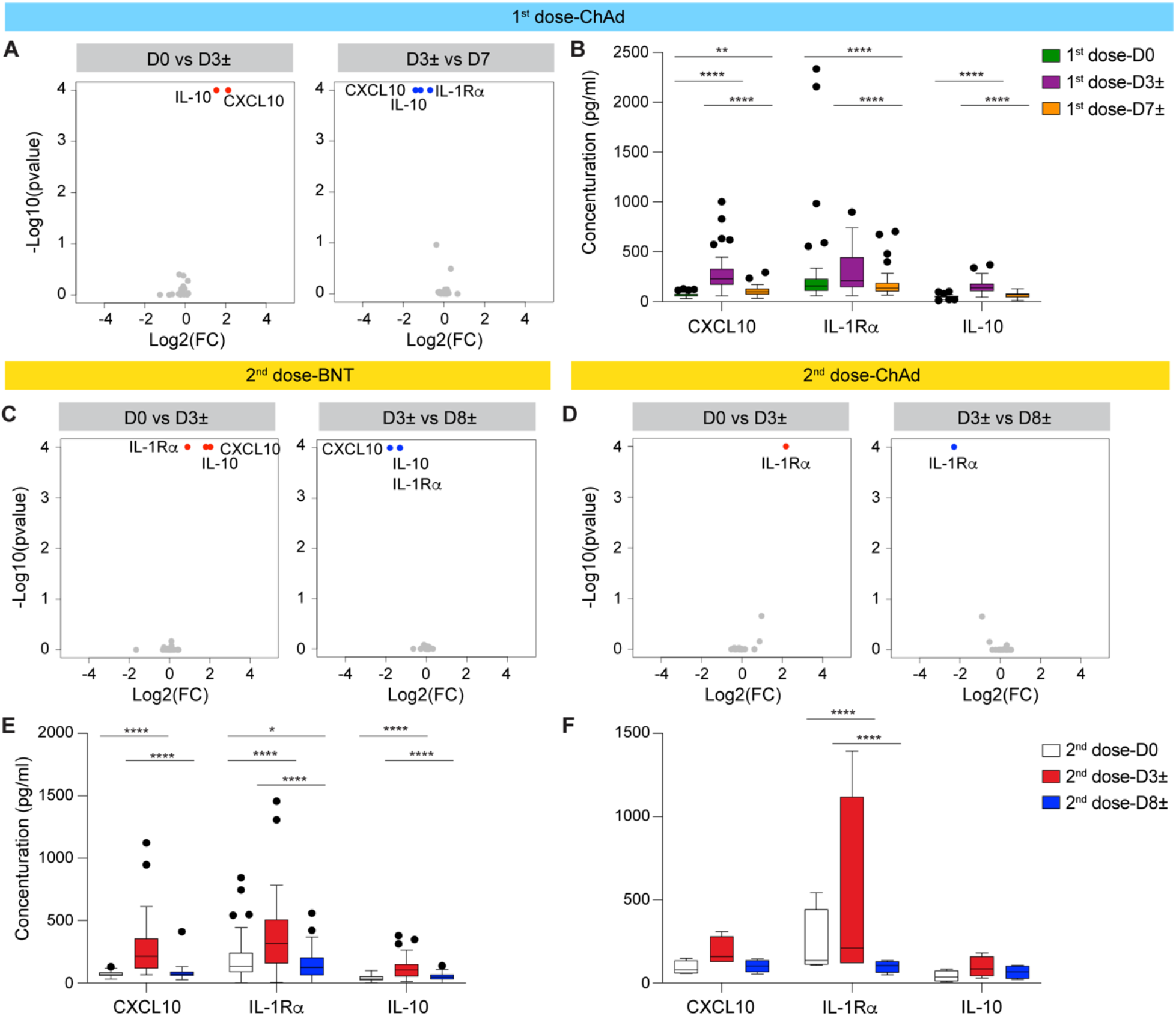
Serum cytokine and chemokine levels. **A** Volcano plots depict differentially expressed cytokines and chemokine at days 0, 3± and 7± of first dose vaccination. Red dots indicate significant upregulation; blue dots indicate significant downregulation (p-value < 0.05). Grey dots indicate cytokines not regulated by the vaccination. **B** CXCL10, IL1-Rα and IL-10 serum concentrations prior to and post vaccination (pg/ml). **C-D** Volcano plots depict differentially expressed cytokines and chemokine at days 0, 3± and 8± after 2nd dose vaccination with either BNT or ChAd. **E-F** Concentrations of CXCL10, IL1-Rα and IL-10 in serum after second vaccine dose. Boxes show median, 25th and 75th percentiles and whiskers show the range. *p*-value are from 2-way ANOVA with Tukey’s multiple comparisons test and listed in Supplementary Table 3. \**p* < 0.05, \*\**p* < 0.01, \*\*\*\**p* < 0.0001. Median, middle bar inside the box; IQR, 50% of the data; whiskers, 1.5 times the IQR.

### Transcriptome response

To further understand the molecular underpinnings of the stark differences in antibody responses observed in the two cohorts after the second dose, BNT versus ChAd, we analyzed the transcriptomes induced after the first and second vaccine dose (Figure 4; Supplementary Figure 2; Supplementary Tables 4-14). Expression of 308 genes was activated within two days (D2) after the prime ChAd vaccination (Figure 4A; Supplementary Table 4). GSEA analyses linked them to innate immune responses including interferon and cytokine signaling (Figure 4D). Expression of 132 genes remained elevated at day 7 (Figure 4A; Supplementary Table 7).

**Figure 4.**
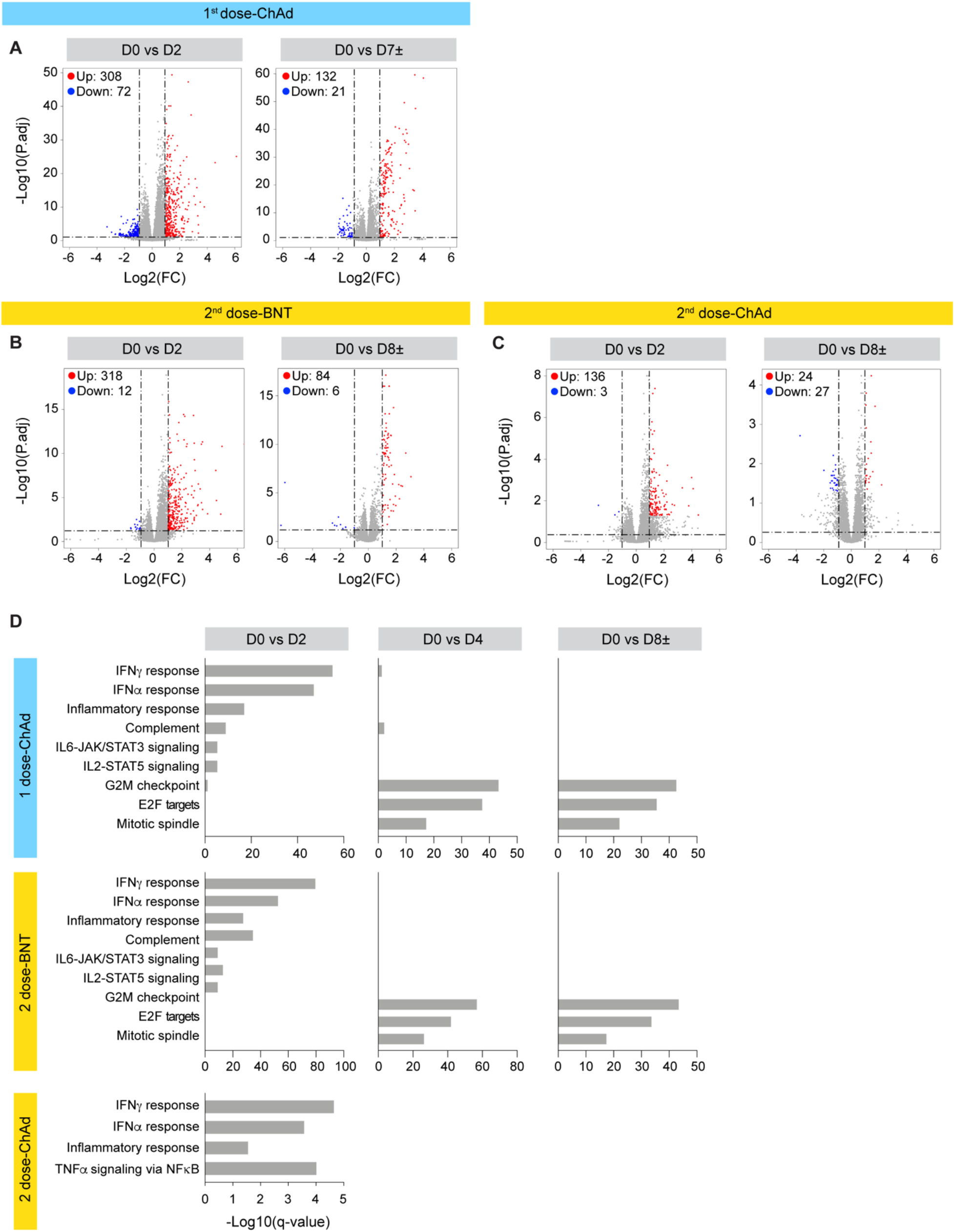
Immune transcriptomes following vaccination. **A** Volcano plots of DEGs comparing Day0 versus Day2 and Day0 vs Day7± in the 1^st^ dose vaccination. DEGs (adjusted p-value, P.adj < 0.05) with a log2 fold change (FC) of more than 1 or less than -1 are indicated in red and blue, respectively. Non-significant DEGs are indicated in gray. The numbers of upregulated and downregulated genes are listed in Supplementary Tables 4-7. **B-C** Volcano plot of DEGs comparing Day0 versus Day2 and Day0 vs Day8±. The numbers of upregulated and downregulated genes are listed in Supplementary Tables 8-13. **D** Genes expressed at significantly higher levels between days were significantly enriched in Hallmark gene sets. X-axis denotes statistical significance as measured by minus logarithm of FDR q-values. Y-axis ranked the terms by q values.

The transcriptome induced by BNT (heterologous group) distinctly differed from that induced by ChAd (homologous group). While 318 genes were significantly activated upon BNT vaccination, ChAd elicited the induction of 136 genes (Figure 4B-D; Supplementary Tables 8 and 12). The 318 BNT-induced genes are enriched in IFN response, complement and cytokine signaling, and the 136 ChAd-induced genes are associated with IFN response and TNF signaling. Of them, 29 genes overlapped in both cohorts, and they are related to IFN responses (Supplementary Figure 2; Supplementary Table 14). Most genes that were activated at day 7 following the BNT vaccination are associated with proliferative responses. In contrast, the 24 genes induced after the second ChAd vaccination are not associated with any GSEA gene category (Figure 4C-D). These findings indicate a prolonged vaccine-induced transcriptomic response in the heterologous ChAd-BNT group as compared to the homologous ChAd-ChAd group.

Recently, we investigated the molecular immune response to a homologous BNT-BNT vaccine protocol in a cohort (n=14, median 58 years old) in Austria^15^. We now compared the immune response elicited by the BNT-BNT and the ChAd-BNT regimen after the first and second dose. While 132 genes were induced by the prime ChAd dose, 75 were induced by the prime BNT vaccination, with seven cell cycle associated genes induced by both vaccines (Supplementary Figure 3; Supplementary Table 14). Following the second vaccination, 83 genes that are enriched in IFN responses, inflammation, complement and cytokine signaling were induced by BNT in both cohorts (Supplementary Figure 4A-B; Supplementary Table 14). Of them six were also identified in the ChAd-ChAd cohort after the second dose (Supplementary Figure 4C; Supplementary Table 14).

### Germline allelic variants of immunoglobulin loci

The preferential increase in anti-spike antibodies and neutralizing antibodies in the heterologous cohort upon receiving the second (BNT) vaccine, led us to dig deeper and interrogate the expression profiles of specific germline variable gene classes. Our sequencing depth exceeded 240 million reads per sample and therefore permitted a detailed analysis of germline gene usage. We determined the range of IGHV, IGKV and IGLV gene usage in the ChAd-BNT heterologous and ChAd-ChAd homologous group after the second vaccination (Figure 5; Supplementary Figures 5-6; Supplementary Table 15). RNA-seq was conducted prior to the first and second vaccination (D0) and eight days (D8) after the second dose. Analysis of immunoglobulin G heavy chain variable (IGHV) and light chain variable (IGKV and IGLV) genes with first complementarity determining region (CDR1) and CDR2 revealed the use of a broad range of germlines in both cohorts and the increase of the total number of clones after the second dose was significant in the ChAd-BNT group (Figure 5A; Supplementary Table 15). We observed increased transcription of several VH genes, including IGHV3-30, IGHV1-18, IGHV1-69 and IGHV3-53 in the heterologous ChAd-BNT group and to a lesser extent in the ChAd-ChAd group (Figure 5B; Supplementary Figure 5A; Supplementary Table 15). These germline allelic variants are the basis of neutralizing antibodies, targeting the spike protein receptor binding domain (RBD), produced after SARS-CoV-2 infection^20-23^. Similarly, specific IGKV and IGLV allelic variants were preferentially induced in the heterologous vaccine group and some variants were induced in both groups (Figure 5C-D; Supplementary Figure 5B-C; Supplementary Table 15).

**Figure 5.**
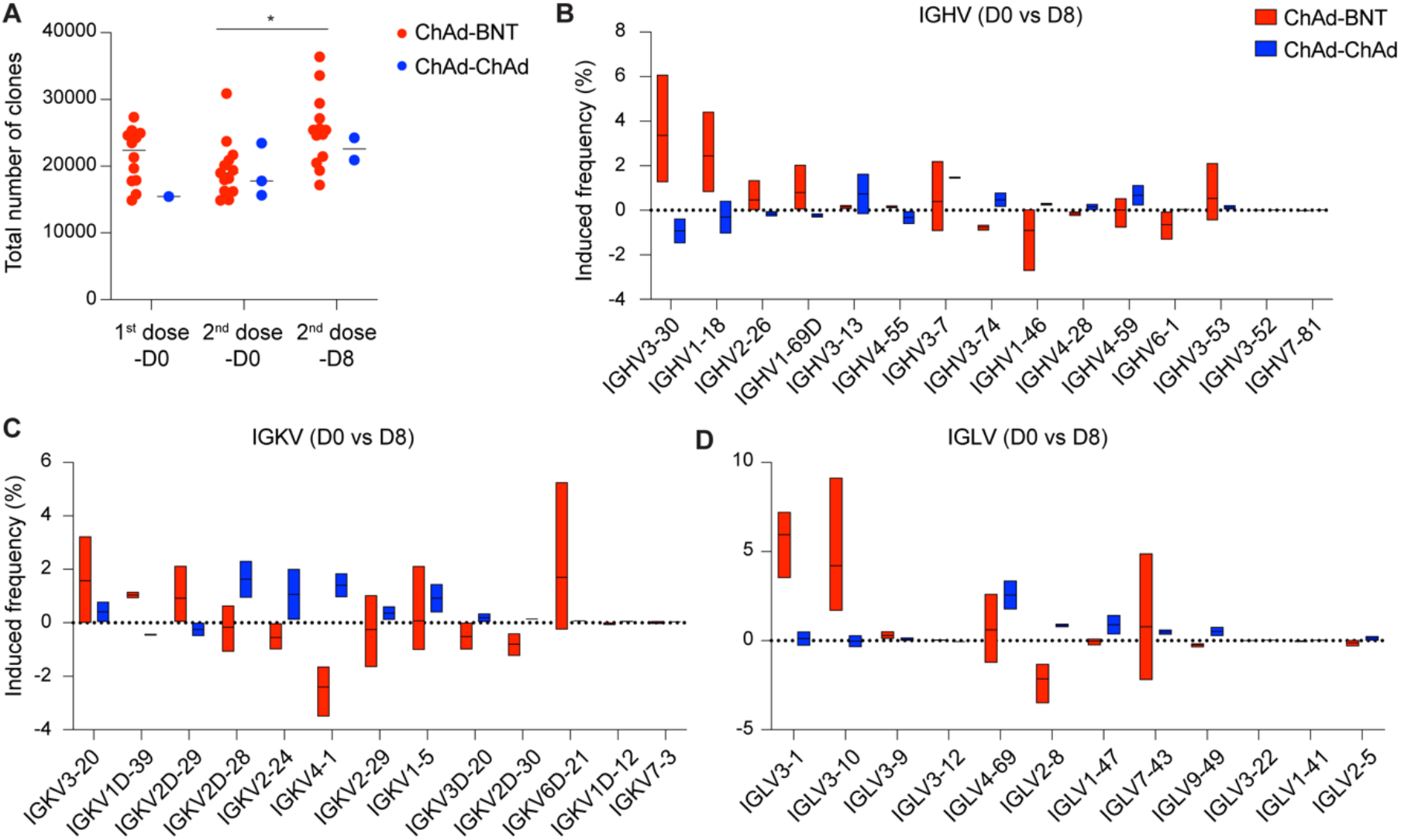
SARS-CoV-2-RBD-specific B-cell memory. **A** The total number of immunoglobulin genes that were expressed at different time points prior to and post vaccination. **B-D** Induction frequency of specific variable gene classes for all paired heavy (B), kappa (C), and lambda (D) chains identified in the three ChAd-BNT and two BNT-BNT individuals prior to the vaccination and after 7 days (Supplementary Table 14). Floating bars (min to max); line at mean.

Next, we examined the rearrangement of the three gene segments (IGV-IGD-IGJ) that generate CDR 3 regions, and CDR3 sequences (Supplementary Figure 6; Supplementary Table 15). Specifically, IGHD3-10, IGHD1-26, IGHD6-13 were highly induced in the heterologous cohort (Supplementary Figure 6A). A similar pattern was observed for the level of J genes (IGHJ, IGKJ, IGLJ) and CDR3 (Supplementary Figure 6B-D).

In a quest to understand whether the ChAd-BNT and BNT-BNT vaccine regimen elicited distinctly different germline allelic variants, we integrated data from this study with our previous vaccination study in Tyrol^15^ (Supplementary Figure 7; Supplementary Table 16). The number of clones in the homologous BNT-BNT cohort increased almost two-fold after the second dose and approximately 1.3-fold in the ChAd-BNT heterologous cohort (Supplementary Figure 7A; Supplementary Table 16). IGHV3-30 and IGHV4-61 clones, the basis of neutralizing antibodies against SARS-CoV-2 infection^24,25^, were induced in both groups, yet their levels were higher in the BNT-BNT group (Supplementary Figure 7B). IGHV2-70 that was detected in Korean COVID-19 patients as a potently therapeutic neutralizing antibody^26^ was induced by BNT, but not ChAd, in the ChAd-BNT and BNT-BNT group after 2nd dose (Supplementary Tables 15-16). However, more diverse immunoglobulin genes were induced in the BNT-BNT group compared to the ChAd-BNT group. A similar pattern was observed for IGKV and IGLV genes (Supplementary Figure 7C-D). In contrast to BCR genes, the activation of TCR genes (TRAV, TRBV) was activated higher in the ChAd-BNT group compared to the BNT-BNT group (Supplementary Figure 7E-F; Supplementary Table 17). T-cell responses are greater in the heterologous group (ChAd-BNT) compared with the homologous group (BNT-BNT, ChAd-ChAd)^27^.

Single cell RNA-seq (scRNA-seq) and CyTOF data of PBMC or whole blood from COVID-19 infection and SARS-CoV-2 mRNA vaccination reveals an increase of monocytes, B cells and CD4+ T cells^9,28-30^. To specifically assess distinct memory response induced by the heterologous and homologous vaccinations, we performed scRNA-seq and identified cell populations in PBMC. B cell populations, especially memory B cells, were elevated in the ChAd-BNT group while T cell populations were higher in the ChAd-ChAd group at day 8 after the second dose (Figure 6). COVID-19 disease severity is related to comparably activation of B cells and substantially reduced T cells^30^.

**Figure 6.**
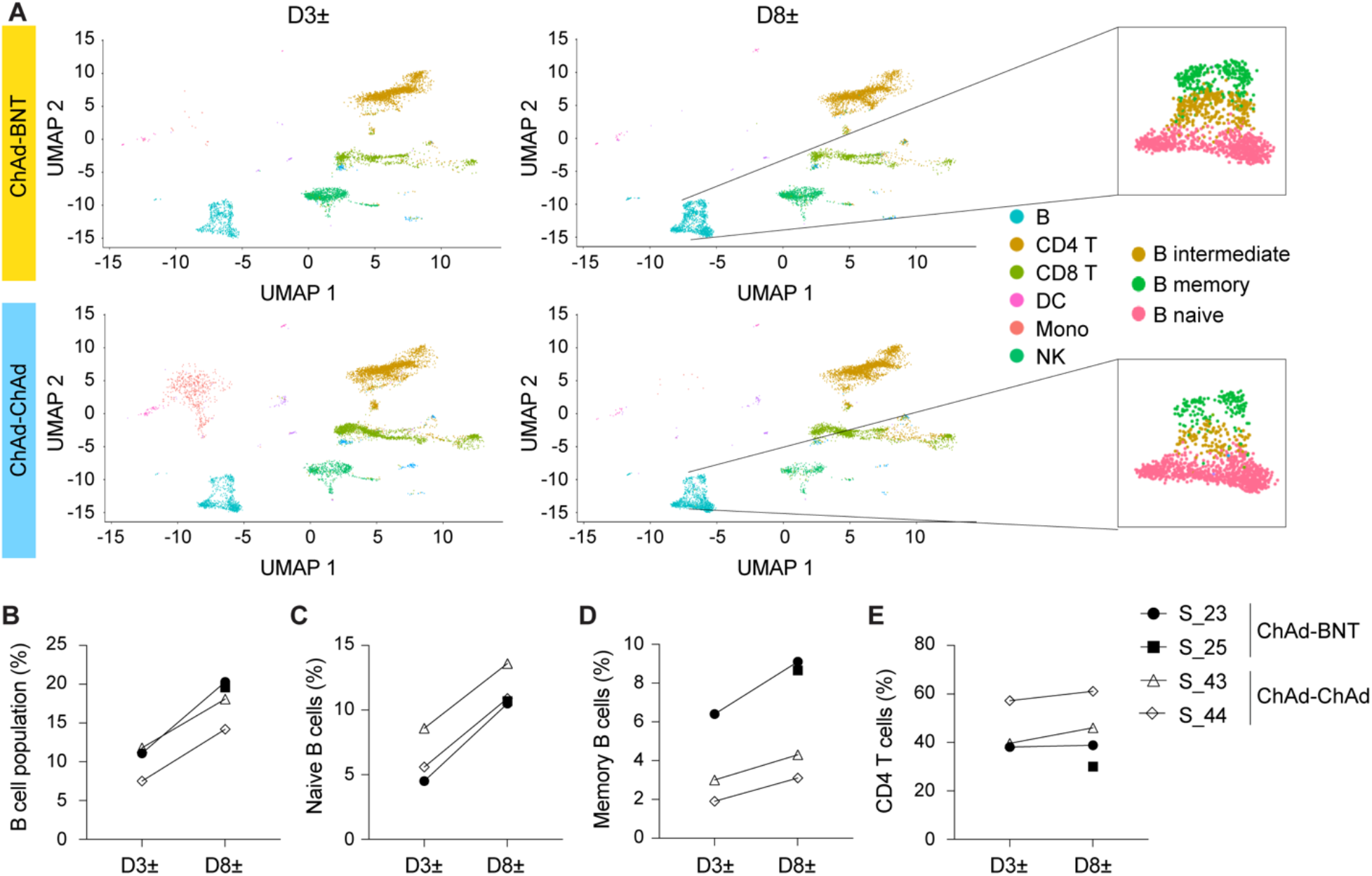
Dissection of immune dynamics using scRNA-seq. **A** Feature plots across time points in the ChAd-BNT and ChAd-ChAd groups. The UMAP projection of PBMCs was annotated by timepoints and colored by individuals. **B-E** Frequencies of total B, naïve B, memory B and CD4+ T cells following vaccination.

## Discussion

Our study adds to a molecular understanding of findings that a heterologous ChAdOx1-BNT162b2 vaccination schedule promotes a more robust immune response than two doses of either BNT162b or ChAdOx1^2-4^. While several ChAd-ChAd heterologous vaccination studies^5,6^, including from South Korea^11,31^, have been published, this is the first report investigating the molecular immune response using bulk and single cell RNA-seq as well as the antibody response to different variants, including the fast-spreading Omicron.

Surveying vaccine-induced genomic responses in peripheral immune cells through RNA-seq approaches aids the identification of transcriptional signatures associated with effective antibody production^9,21,32^. Our sequencing depth of more than 240 million reads per sample permitted the identification of gene signatures linked to either the heterologous ChAd-BNT or the homologous ChAd-ChAd group. The BNT vaccine in the heterologous group elicited a stronger interferon signature with an activated JAK-STAT pathway than the second ChAd dose or the second BNT dose in the homologous group. Our analysis permitted the preferential activation of specific variable germline classes as well as CDR3 classes in the ChAd-BNT and ChAd-ChAd groups. An increase of specific IGHV clonal transcripts encoding neutralizing antibodies was preferentially detected in the heterologous cohort and the homologous BNT-BNT cohort.

The benefits of adding BNT162b2 as the second dose in heterologous vaccination regimens with ChAdOx1 as the primary dose is well established^5,33^. In extension, recent studies demonstrated that a three-dose heterologous regimen with two doses of CoronoVac immunization followed by BNT162b2 was associated with a 1.4-fold increase of neutralizing antibodies against Omicron as compared to a homologous two-dose BNT162b2 regimen^34,35^. However, neutralizing activity against Omicron was reduced by 7.1-fold and 3.6-fold compared to the ancestral and Delta variants, respectively. Here we show that a two-dose heterologous vaccine regimen with initial ChAd vaccination followed by BNT resulted in Omicron targeted antibody titers less than 3.2-fold reduced than that found against the ancestral variant as compared to 5.9-fold reduction with ChAd followed by ChAd, and a 4.7-fold reduction in a historical cohort receiving BNT-BNT. Further evolution of COVID-19 variants is an open question, but it is likely that additional variants will emerge, with manufacture of targeted vaccines following their identification. Therefore, it is reasonable to examine vaccine regimens for their ability to induce a range of antibody response that may provide coverage for emerging variants. Here the ChAd-BNT regimen appeared to be successful for that purpose. To date, up to 2.5 billion doses of ChAd have been delivered with an additional 500 million doses of alternative adenovirus based vaccines from Johnson and Johnson and Sputnik from Russia^36^. Data presented here supports that from an immunological perspective of a heterologous adenovirus-based mRNA-based regimen is effective and may be a reasonable cost-effective approach for specific populations where public health officials deem that adenovirus-based vaccine side effects are low risk. A heterologous strategy might also foster enhanced immune responses in immunocompromised individuals.

### Limitations of the study

There are several limitations to this study. First, our study population was limited to a specific geographic area (South Korea) and a specific genetic population. Second, most study subjects were healthy females of normal weight. Third, the homologous ChAd-ChAd cohort was smaller than the heterologous ChAd-BNT cohort. Fourth, our study did not investigate neutralizing antibodies using live or pseudo-SARS-CoV-2 virus and variants.

## Methods

### Study population, study design and recruitment

51 SARS-CoV-2 naïve healthy volunteers (Supplementary Table 1) were recruited for the study under informed consent. Recruitment and blood sample collection took place between June and September 2021. This study was approved (IRB No 2021-0898) by the Institutional Review Board (IRB) of Asan medical center in Korea. Written informed consent was obtained from all subjects. This study was determined to impose minimal risk on participants. All methods were carried out in accordance with relevant guidelines and regulations. All research has been have been performed in accordance with the <u>Declaration of Helsinki</u> (https://www.wma.net/policies-post/wma-declaration-of-helsinki-ethical-principles-for-medical-research-involving-human-subjects/). In addition, we followed the ‘<u>Sex and Gender Equity in Research – SAGER – guidelines</u>’ and included sex and gender considerations where relevant.

### Quantification of immunoproteins

Plasma samples from all participants were collected from their blood. After thawing, the samples were centrifuged for 3 minutes at 2000 *g* to remove particulates prior to sample preparation and analysis. Human XL Cytokine Luminex Performance Panel Premixed Kit (R&D systems, #FCSTM18) was used to measure proinflammatory proteins (IFN-α, IFN-*γ*, IL-1α, IL-1β, IL-1Rα, IL-2, IL-3, IL-4, IL-6, IL-7, IL-8, IL-10 and TNF-α), cytokines (IL-12p70, IL-13, IL-15, IL1-7A and VEGF) and chemokine (CCL2, CCL3, CCL4, CCL11 and CXCL10). The analysis was performed according to the manufacturers’ protocols.

### COVID-19 serology and ACE2-neutralization assay

A multiplexed solid-phase chemiluminescence assay (Meso Scale Discovery, MD) was evaluated for the detection of IgG binding to various SARS-CoV-2–derived antigens (V-PLEX SARS-CoV-2 Panel 17 (IgG) Kit, K15524U, and Panel 23 (IgG) Kit, K15567U) and the quantification of antibody-induced ACE-2 binding inhibition to various variants’ spike antigens (pseudo-neutralization assay) (V-PLEX SARS-CoV-2 Panel 18 (ACE2) Kit, K15535U, and Panel 23 (ACE2) Kit, K15570U). Plates were coated with the specific antigen on spots in the 96 well plate and the bound antibodies in the samples were then detected by anti-human IgG antibodies or ACE2 conjugated with the MSD SULPHO-TAG which is then read on the MSD instrument which measures the light emitted from the tag.

### Extraction of the buffy coat and purification of RNA

Whole blood was collected, and total RNA was extracted from the buffy coat and purified using the RNeasy Mini Kit (Qiagen, #74104) according to the manufacturer’s instructions. The concentration and quality of RNA were assessed by an Agilent Bioanalyzer 2100 (Agilent Technologies, CA).

### mRNA sequencing (mRNA-seq) and data analysis

The Poly-A containing mRNA was purified by poly-T oligo hybridization from 1 mg of total RNA and cDNA was synthesized using SuperScript III (Invitrogen, MA). Libraries for sequencing were prepared according to the manufacturer’s instructions with TruSeq Stranded mRNA Library Prep Kit (Illumina, CA, RS-20020595) and paired-end sequencing was done with a NovaSeq 6000 instrument (Illumina) yielding 200-350 million reads per sample.

The raw data were subjected to QC analyses using the FastQC tool (version 0.11.9) (https://www.bioinformatics.babraham.ac.uk/projects/fastqc/). mRNA-seq read quality control was done using Trimmomatic^37^ (version 0.36) and STAR RNA-seq^38^ (version STAR 2.5.4a) using 150 bp paired-end mode was used to align the reads (hg19). HTSeq^39^ (version 0.9.1) was to retrieve the raw counts and subsequently, Bioconductor package DESeq2^40^ in R (https://www.R-project.org/) was used to normalize the counts across samples and perform differential expression gene analysis. Additionally, the RUVSeq^41^ package was applied to remove confounding factors. The data were pre-filtered keeping only genes with at least ten reads in total. The visualization was done using dplyr (https://CRAN.R-project.org/package=dplyr) and ggplot2^42^. The genes with log2 fold change >1 or <-1 and adjusted p-value (pAdj) <0.05 corrected for multiple testing using the Benjamini-Hochberg method were considered significant and then conducted gene enrichment analysis (GSEA, https://www.gsea-msigdb.org/gsea/msigdb).

For T- or B-cell receptor repertoire sequencing analysis, trimmed fastq files from bulk RNA-seq were aligned against human V, D and J gene sequences using the default settings with MiXCR^43,44^. CDR3 sequence and the rearranged BCR/TCR genes were identified.

### Single-cell RNA sequencing (scRNA-seq) and data analysis

The isolated PBMCs were frozen in freezing media (Thermofisher) and stored at -80°C until use. Single-cell suspensions were then immediately loaded on the 10X Genomics Chromium Controller with a loading target of 20,000 cells. Libraries were generated using the Chromium Next GEM Single Cell 3′ Kit v3.1 (Dual Index) according to the manufacturer’s instructions. Libraries were sequenced using the NovaSeq 6000 instrument (Illumina).

The raw reads were aligned and quantified using the Cell Ranger with Feature Barcode addition (version 5.0, 10X Genomics) against the GRCh38 human reference genome. The quality control, normalization, dimension reduction, cell clusters, UMAP projection, and cell type annotation were performed using Seurat (version 4.0)^45^.

### Statistical analysis

Differential expression gene (DEG) identification used Bioconductor package DESeq2 in R. P-values were calculated using a paired, two-side Wilcoxon test and adjusted p-value (pAdj) corrected using the Benjamini–Hochberg method. Genes with log2 fold change >1 or <-1, pAdj <0.05 and without 0 value from all sample were considered significant. For significance of each GSEA category, significantly regulated gene sets were evaluated with the Kolmogorov-Smirnov statistic. *P*-values of cytokines were calculated using two-stage linear step-up procedure of Benjamini, Krieger and Yekutieli on GraphPad Prism software (version 9.0.0). For comparison of RNA expression levels, antibody levels or cytokine levels between two groups, data were presented as standard deviation in each group and were evaluated with a two-way ANOVA followed by Tukey’s multiple comparisons test using GraphPad PRISM (version 9.0). A value of \**P* < 0.05, \*\**P* < 0.01, \*\*\**P* < 0.001, \*\*\*\**P* < 0.0001 was considered statistically significant.

### Ethics statement

This study was approved by the Institutional Review Board (IRB) of Asan medical center in Korea (IRB number 2021-0898). Participant information was coded and anonymized.

### Data availability

The RNA-seq and scRNA-seq data from this study will be uploaded in GEO before publishing the manuscript.

## Acknowledgments

This work was supported by the Intramural Research Programs (IRPs) of the National Institute of Diabetes and Digestive and Kidney Diseases (NIDDK) and by a grant of the Korea Health Technology R&D Project through the Korea Health Industry Development Institute (KHIDI), funded by the Ministry of Health & Welfare, Republic of Korea (grant number: HI18C2383).

Our gratitude goes to the participants who contributed to this study to advance our understanding of COVID-19 vaccination. We thank Yuhai Dai from the NIDDK clinical core for helping antibody assay. This work was utilized the computational resources of the NIH HPC Biowulf cluster (http://hpc.nih.gov). RNA-sequencing and single cell RNA-sequencing was conducted in the NIH Intramural Sequencing Center, NISC (https://www.nisc.nih.gov/contact.htm) and the NHLBI DNA sequencing and genomics core (https://www.nhlbi.nih.gov/science/dna-sequencing-and-genomics-core), respectively.

## Author contributions

H.K.L., L.H. and J.W.H. designed the study. J.W.H. recruited participants and collected material. HSS collected material and performed cytokine and antibody assays. J.Y.K and S.W.K isolated PBMC and RNA. H.K.L. analyzed RNA-seq and scRNA-seq data. J.W.H. conducted immunoproteins assay. M.W. conducted IgG antibody and neutralization assay. H.K.L., P.A.F., L.H. and J.W.H. analyzed data. H.K.L. administrated project. L.H. and J.W.H. supervised project. H.K.L., P.A.F., L.H. and J.W.H. wrote the paper. All authors read and approved the manuscript.

